# Endemic Seasonal Coronavirus Neutralisation and COVID-19 severity

**DOI:** 10.1101/2021.09.29.21264328

**Authors:** David A. Wells, Diego Cantoni, Martin Mayora-Neto, Cecilia Di Genova, Alexander Sampson, Matteo Ferrari, George Carnell, Angalee Nadesalingam, Peter Smith, Andrew Chan, Gianmarco Raddi, Javier Castillo-Olivares, Helen Baxendale, Nigel Temperton, Jonathan L. Heeney

## Abstract

The virus SARS-CoV-2, responsible for the global COVID-19 pandemic, spread rapidly around the world causing high morbidity and mortality because humans have no pre-existing immunity. However, there are four known, endemic seasonal coronaviruses in humans (HCoVs) and whether antibodies for these HCoVs play a role in severity of COVID-19 disease has generated a lot of interest. Of these seasonal viruses NL63 is of particular interest as it uses the same cell entry receptor as SARS-CoV-2.We use functional, neutralising assays to investigate cross reactive antibodies and their relationship with COVID-19 severity. We analysed neutralisation of SARS-CoV-2, NL63, HKU1, and 229E in 38 COVID-19 patients and 62 healthcare workers, and a further 182 samples to specifically study the relationship between SARS-CoV-2 and NL63.We found that although HCoV neutralisation was very common there was little evidence that these antibodies neutralised SARS-CoV-2. Despite no evidence in cross neutralisation, levels of NL63 neutralisating antibodies become elevated after exposure to SARS-CoV-2 through infection or following vaccination.

## Introduction

The human endemic coronaviruses (HCoVs), sometimes referred to as seasonal coronaviruses, are a group of four viruses that include the alphacoronaviruses 229E, NL63, and two betacoronaviruses HKU1, OC43. Frequently infecting humans through life and generally causing symptoms of a common cold (1), they are considered to be low morbidity pathogens. On rare occasions however, infection in individuals with serious underlying diseases can lead to severe pneumonia and death (2,3). The HCoVs are known to have a wide tropism, however there are selective binding profiles to certain sugars and proteins. HKU-1 and OC43 bind to sialic acid (4), 229E binds to human aminopeptidase N (5) and NL63 binds to the angiotensin-converting enzyme 2 (ACE2) receptor (6). The severe acute respiratory syndrome coronavirus 2 (SARS-CoV-2) has been rapidly transmitted and spread globally, causing over 200 million cases and more than 4 million deaths as of September, 2021. Previous studies have demonstrated pre-existing immune responses to SARS-CoV-2 in people not exposed to the virus. This has been reported both for antibodies (7–9), and T cell responses (10,11). Though still under debate (12,13) this pre-existing immunity to a novel virus has largely been attributed to the four widely circulating HCoVs.

There has been great interest in a potential role of common cold coronaviruses modulating the severity of COVID-19 disease. This is partly due to the fact that NL63 also uses the same ACE2 as its cellular receptor; therefore, it was questioned whether antibodies raised against NL63 would also bind to SARS-CoV-2. A recent report had investigated whether seasonal HCoVs could protect against SARS-CoV-2 infection (14). Anderson et al. used antibody binding assays (ELISAs) to quantify antibodies against the HCoVs. However, these binding assays do not discriminate between neutralising and non-neutralising antibodies. Pseudotyped viruses (PVs) can be used to quantify neutralising antibodies and have been shown to correlate with live-virus neutralisation (15–17).

Herein, we used PVs bearing the Spike protein of SARS-CoV-2 and the seasonal HCoVs: NL63, HKU1 and 229E to investigate the relationships between common cold coronavirus immune responses and COVID-19 severity in healthcare workers and COVID-19 patients. We found that HCoV neutralisation did not correlate with SARS-CoV-2 neutralisation. This builds on previous work showing that HCoV binding does not protect against COVID-19 (14); however, we also show that HCoV binding and HCoV neutralisation are not strongly correlated. This finding highlights the importance of functional antibody assays, in addition to binding, when characterising antibody responses. Despite a lack of cross neutralisation we found that NL63 neutralisation is boosted by SARS-CoV-2 vaccination, and elevated after moderate to mild COVID-19 disease.

## Methods

### Subject recruitment and serum collection

Health care workers (HCWs) and COVID-19 patients were recruited from Royal Papworth Hospital, Cambridge, United Kingdom in spring 2020. HCW were recruited through staff email over the course of 2 months (20^th^ April 2020-10^th^ June 2020) as part of a prospective study to establish seroprevalence and immune correlates of protective immunity to SARS-CoV-2. Following informed consent, staff were invited to complete a questionnaire to clarify whether they had swab PCR confirmed SARS-CoV-2 infection (routine swabbing was not available at that time and there was limited access to swabbing when symptomatic) and whether they had experienced symptoms that they felt may have been consistent with COVID-19 since January 2020. Symptom severity was classified according to WHO severity classification into asymptomatic, mild, moderate and severe disease. The study was approved by Research Ethics Committee Wales, IRAS 96194 12/WA/0148. Amendment 5. All participants provided written and informed consent prior to being enrolled in this study.

Serum was taken from HCWs and convalescent COVID-19 patients 0-6 months after recruitment. We analysed SARS-CoV-2 neutralisation and HCoV neutralisation for 38 COVID-19 patients, 23 seropositive HCWs, and 39 seronegative HCWs. These samples are described in more detail in. Because of our interest in NL63 analysed a further set of samples for SARS-CoV-2 and NL63 neutralisation: 35 seropositive HCWs, 140 seronegative HCWs, and 7 COVID-19 patients (6 were seropositive). We also collected follow up samples from 21 of our HCWs 1 month after they received their first SARS-CoV-2 vaccination dose, approximately 9-12 months after they were first recruited to our study.

### Classifying sample serostatus

Samples’ serostatus was determined according to SARS-CoV-2 IgG binding status. This was determined by neutralisation (SARS-CoV-2 pMN IC50), and/or IgG binding to SARS-CoV-2 Spike, Nucleocapsid, and Spike receptor binding domain (RBD). These two methods of classification showed good agreement but as neither assay was performed on all samples a positive result on either assay classified the sample as seropositive. The seropositive cutoff for pMN was the 95% upper confidence interval of pre-pandemic samples in previous work (18). The classification based on IgG binding is described in (19) but in brief a linear support vector machine was trained to distinguish a set of pre-pandemic sera from COVID-19 patient sera. This classification method considers the three antigens jointly so there is no single cut-off to report.

### Tissue Culture

Human Embryonic Kidney cells (HEK293T/17) cells and Huh-7 based hepatoma cells were maintained using DMEM supplemented with 10% foetal bovine serum (FBS) and 1% penicillin/streptomycin (P/S). Chinese Hamster Ovary (CHO) cells were maintained in Ham’s F-12 medium supplemented with 10% FBS and 1% P/S. All cells were incubated at 37°C and 5% CO_2_. Cells were routinely passaged three times a week to prevent overconfluency.

### Pseudotype virus generation

PV generation was carried out as previously described (20). Plasmids bearing the Spike of either SARS-CoV-2, NL63, HKU1, and 229E in the vector pcDNA3.1+, were mixed with the plasmids p8.91 lentiviral Gag-pol (21) and pCSFLW luciferase reporter gene (22) in Opti-MEM solution. Plasmids were mixed and incubated for 15 minutes with FuGENE-HD transfection reagent, followed by dropwise addition onto HEK293T cells. For HKU1 PVs, 1.5U of exogenous neuraminidase (Sigma) was added 18-24h post transfection. Cells were incubated for 48 hours prior to removal and filtering of the supernatant culture media through 0.45µM cellulose acetate membranes. Aliquots of filtered supernatant were stored at -80 °C. Pseudotypes were titrated in white flat-bottomed 96 well plates by serially diluting 2-fold into DMEM for PVs bearing the SARS-CoV-2 and NL63 spike, or Ham’s F-12 media for PVs bearing the spike of HKU1. Target cells for SARS-CoV-2 and NL63 PVs were HEK293T cells pre-transfected with ACE-2 and TMPRSS-2 (23), and CHO cells were used as target cells for HKU1 PVs. Plates were incubated at 37°C and 5% CO_2_ for 48 hours prior to lysis using Bright-Glo and assaying luciferase reporter gene activity in relative light units (RLU) using a luminometer. PV titres were reported in RLU/ml.

### Pseudotype virus neutralisation (pMN) Assays

pMN assays were carried out as previously described (20). Briefly, serum was mixed with either DMEM or Ham’s F-12 depending on the PV, at an initial 1:40 dilution and serially diluted 2-fold in white flat-bottomed 96-well plates to a final 1:5,120 dilution. PVs were then added to the wells at an input of 5×10^5^ RLU/ml and plates were incubated at 37°C and 5% CO_2_ for one hour. Pre-transfected HEK293T target cells were seeded at 1×10^4^ cells per well in plates containing either SARS-CoV-2 or NL63 PVs, Huh-7 cells were seeded at 1×10^4^ cells per well in plates containing 229E PVs and CHO cells were seeded at 1×10^4^ cells per well in plates containing HKU1 PVs. Plates were incubated at 37°C and 5% CO_2_ for 48 hours prior to lysis using Bright-Glo and assaying luciferase reporter gene activity in relative light units (RLU) using a luminometer. IC_50_ values were calculated for the neutralisation assays based on 4-parameter log-logistic regression dose response curves. These curves were fit using Autoplate (Palmer et al, *under review*) and the R package drc.

### Statistical methods

#### HCoV neutralisation and COVID-19 severity

We used multiple regression to compare HCoV neutralisation titres between SARS-CoV-2 seropositive HCWs and COVID-19 patients after accounting for differences in age and sex. Age and sex effects were reported after dropping non-significan HCW/patient terms. A linear model predicting HCoV neutralisation was fit separately for NL63, HKU1, and 229E. All statistical analyses were performed using R (24).

We fit a linear regression to predict COVID-19 severity in 81 seropositive people. This model included the natural log of the SARS-CoV-2 pMN IC50 and a binary term indicating whether or not the sample came from a hospitalised COVID-19 patient. We used an F ratio test to determine if the natural log of the NL63 pMN IC50 significantly improved the model fit. Plots of residuals, leverage, and qq-plots were used to assess the assumptions of the model.

The WHO COVID-19 severity score is ordinal so we also analysed our data using a proportional odds logistic regression designed for ordinal variables to ensure our conclusions are robust to non-linearity in the data. Similar to the linear regression, our proportional odds logistic regression predicted COVID-19 severity in 81 seropositive people using the natural log of SARS-CoV-2 pMN IC50 and whether or not the sample came from a COVID-19 patient as predictors. We used a likelihood ratio test to test if the natural log of NL63 pMN IC50 significantly predicted COVID-19 severity after accounting for the other variables. The assumption of proportional odds was assessed by visualising coefficients of logistic regression models predicting severity equal to, or greater than *i* for *i* equals 2-7.

#### Does SARS-CoV-2 exposure increase HCoV neutralisation?

If SARS-CoV-2 infection increased HCoV antibody titre we would expect HCoV neutralisation to be higher in seropositive samples. We compared HCoV neutralisation between serostatus groups to test if SARS-CoV-2 infection increased HCoV neutralisation. We used a linear model using serostatus, sex, and age as predictors.

To investigate the effect of vaccination on NL63 neutralisation we quantified NL63 neutralisation of 21 HCW before and after receiving their first dose of the SARS-CoV-2 vaccination. The significance of any change before and after vaccination was calculated using a paired Wilcoxon signed rank test.

#### Correlations between neutralisation of different viruses and spike binding

We investigated the correlation between neutralisation of SARS-CoV-2 and HCoVs using Spearman’s rank. We also visualised all correlations between HCoV and SARS-CoV-2 neutralisation and spike binding using a Spearman’s rank correlation plot (25).

## Results

### Neutralising antibodies against all three pseudotyped HCoVs detected in plasma samples

To assess if all samples (seronegative HCWs, seropositive HCW, and COVID-19 patients) were positive for seasonal HCoVs we utilised pseudotype virus neutralisation assays. Samples with neutralisation IC50 over 40 were classed as neutralising; the majority of samples neutralised all three HCoVs tested. We found that 98.6% of 282 plasma samples neutralised NL63, 76.4% of 89 samples neutralised HKU1, and 99% of 100 samples neutralised 229E (figure 1). This illustrates the prevalence of HCoV infection and how common it is for people to have circulating neutralising antibodies to HCoVs.

**Figure 1.**
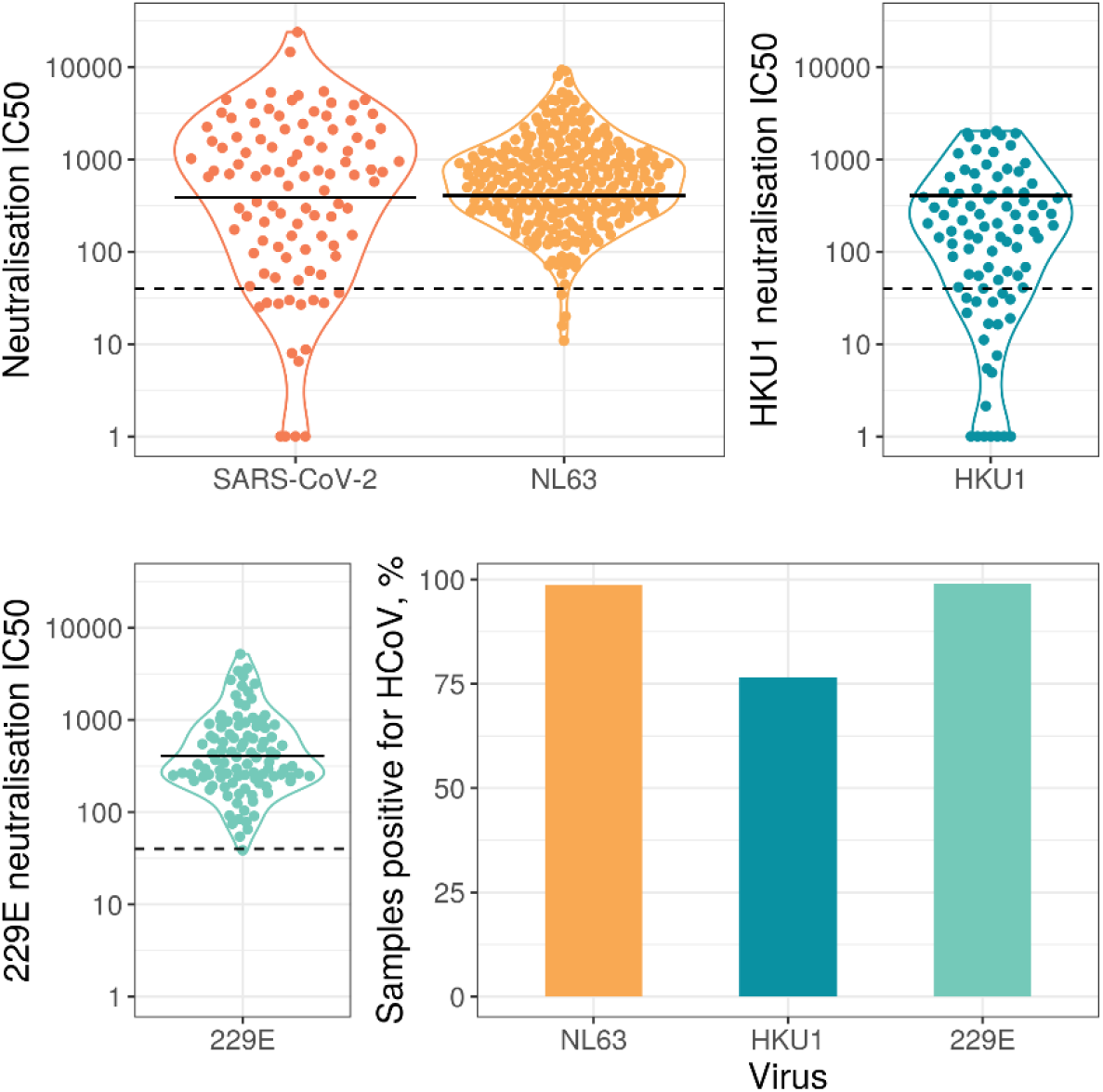
Neutralisation IC50 values for HCoVs and SARS-CoV-2. Solid lines represent geometric means, dashed horizontal lines indicate the cut off chosen to define detectable HCoV neutralisation. Due to the different cell lines used for HKU1 and 229E, data points were plotted on separate graphs. The SARS-CoV-2 data only includes samples from seropositive individuals. Panel d) shows the percentage of samples with detectable HCoV neutralisation, IC50>40.

We found significant sex and age differences in neutralisation titres for some HCoVs when analysing SARS-CoV-2 seropositive samples (figure 2). There was no significant effect for NL63 (sex ß=0.06, SE=0.23, p=0.78; age ß=0.01, SE=0.01, p=0.27). For HKU1 there was a significant effect of sex and age (sex ß=1.9, SE=0.62, p=0.004; age ß=0.05, SE=0.02, p=0.039). For 229E there was a significant effect of age but not sex (sex ß=0.47, SE=0.26, p=0.077, age ß=0.02, SE=0.01, p=0.037).

**Figure 2.**
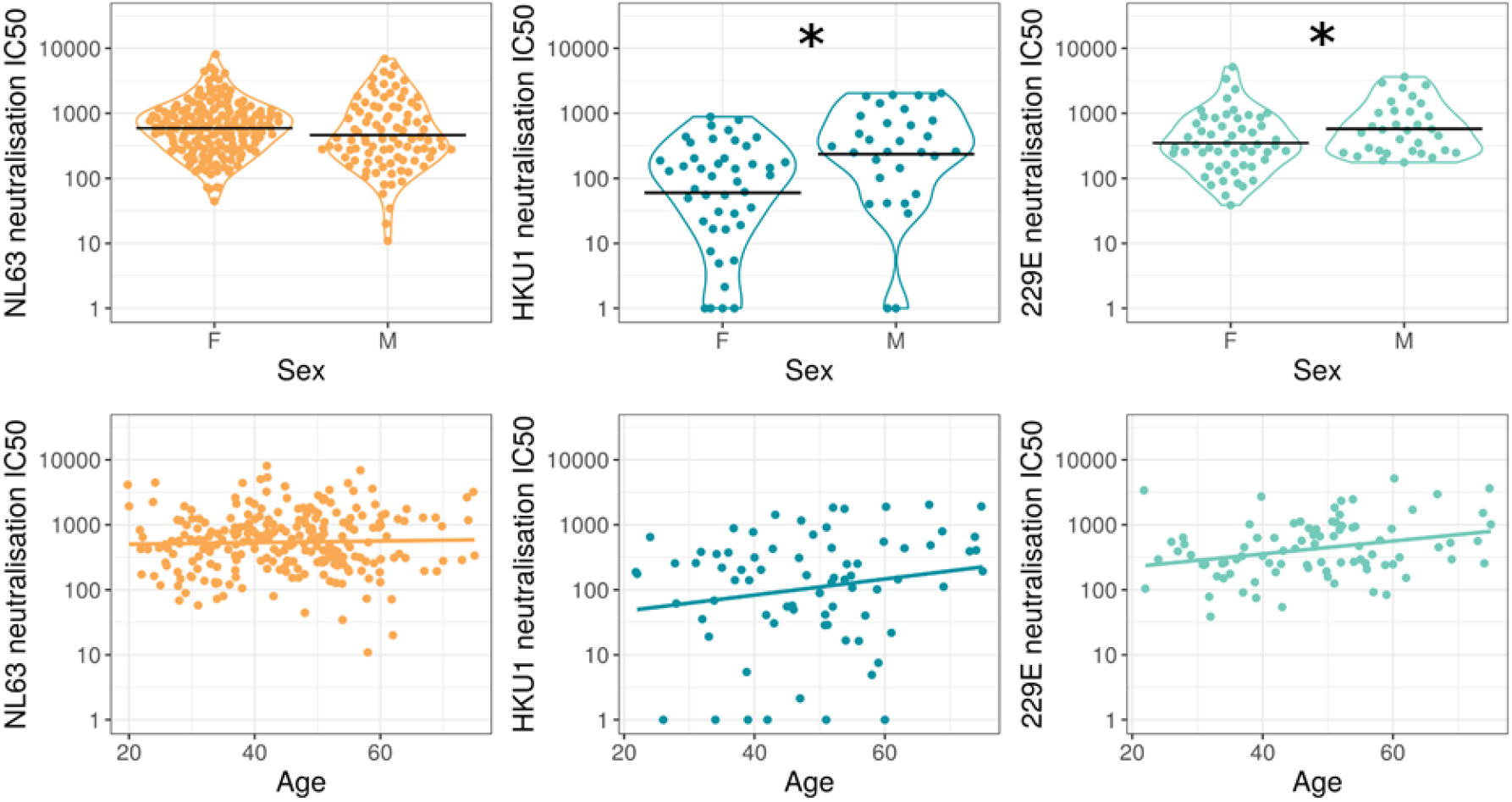
HCoV neutralisation by demographic. Neutralising IC50 value for HCoVs against age and sex. Black horizontal lines represent geometric means, coloured lines in are simple linear regression lines. We found a significant difference in HKU1 neutralisation between sex (p=0.004) and age (p=0.039). We did not see any statistical significance in HCoVs NL63 nor 229E between sexes. We observed a significant difference in 229E neutralisation and age (p=0.037).

### HCoV neutralisation and COVID-19 severity

If HCoV immune responses modulate COVID-19 severity we may observe a difference in HCoV neutralisation between seropositive HCWs and COVID-19 patients. However, after accounting for age and sex we found no significant difference in HCoV neutralisation between SARS-CoV-2 seropositive HCWs and patients (NL63 p=0.150, HKU1 p=0.162, 229E p=0.155, figure 3).

**Figure 3.**
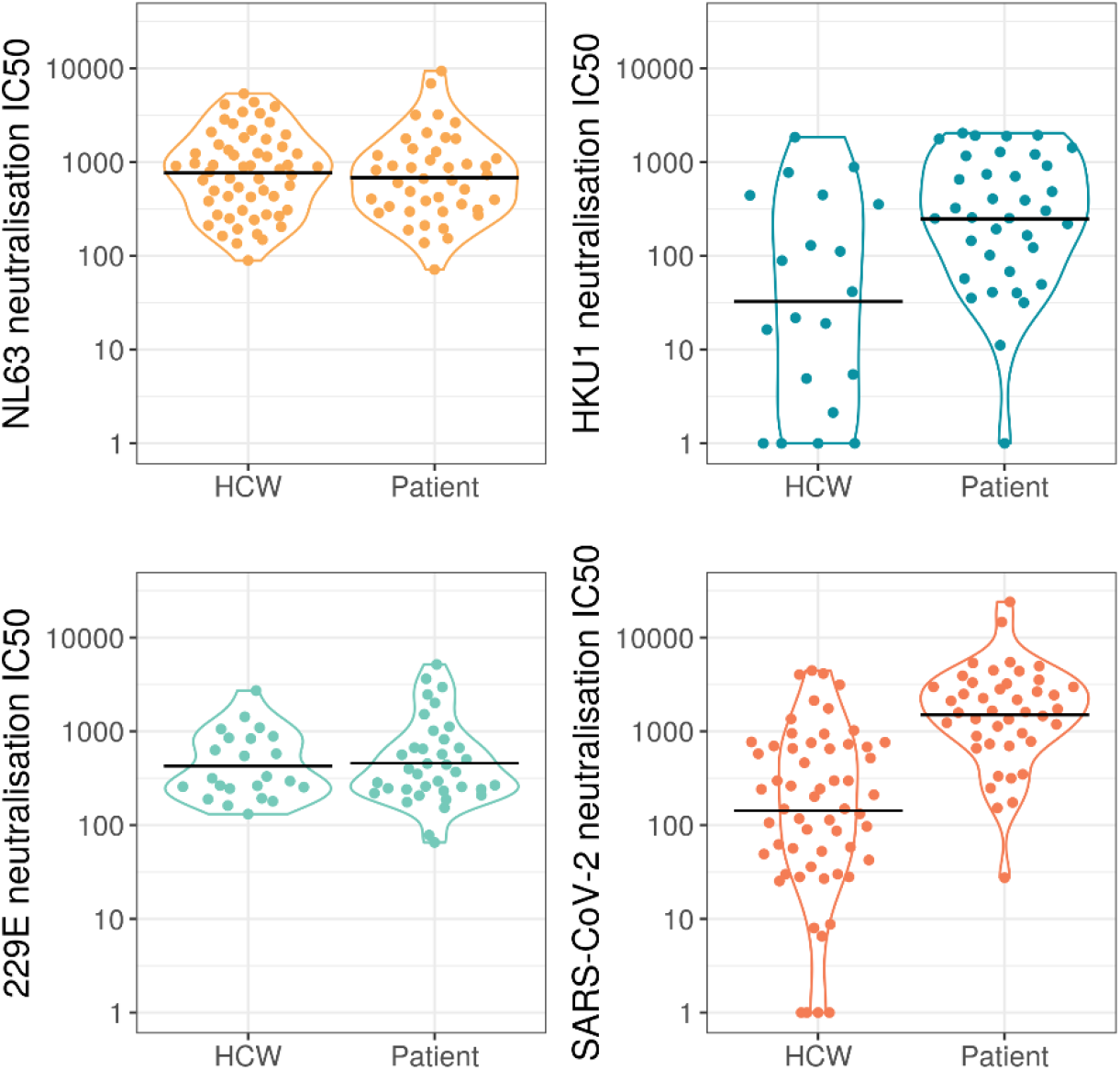
Comparing neutralisation between seropositive HCWs and seropositive patients for NL63 (p=0.150), HKU1 (p=0.162), 229E (p=0.155), and SARS-CoV-2. Horizontal black lines indicate geometric means.

NL63, the only HCoV to use the same ACE2 receptor for cell entry (26), was significantly associated with COVID-19 severity after accounting for SARS-CoV-2 neutralisation (figure 4). This result was found in both our linear regression (F=10.4, DF=1, p=0.002, table 1), and our proportional odds logistic regression (LR=16.7, df=1, p<0.001). Further assessment of the proportional odds logistic regression found that the relationship between NL63 and COVID-19 severity was limited to asymptomatic and very mild cases. This suggests that NL63 neutralisation is elevated in people who have suffered more than very mild COVID-19.

**Figure 4.**
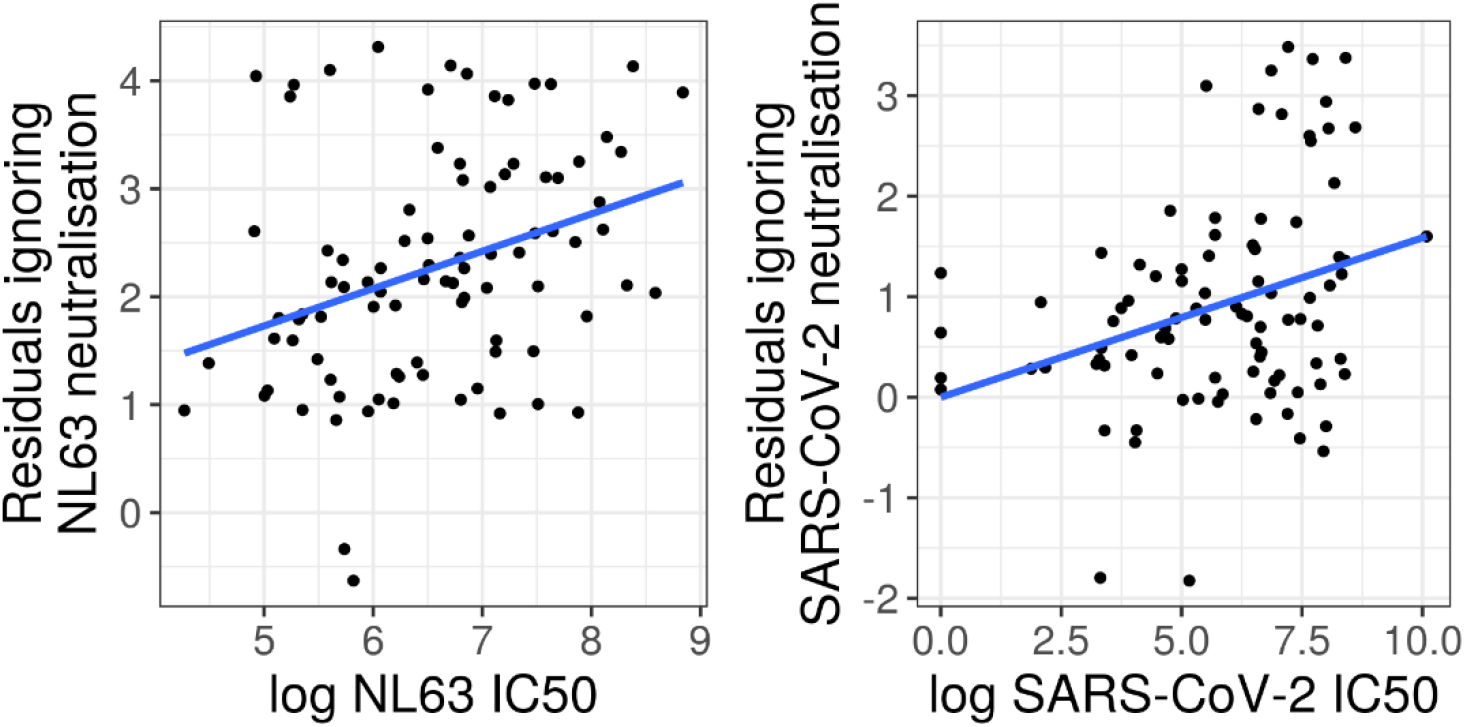
Partial residual plots for the natural log of NL63 and SARS-CoV-2 pMN IC50 values predicting COVID-19 severity. These plots illustrate the effect of a variable on severity after accounting for other variables in the linear regression. This data suggests that NL63 neutralisation is associated with COVID-19 disease severity.

**Table 1.** COVID-19 severity linear regression coefficient estimates and standard errors

| Coefficient | Estimate | Standard Error | p value |
| --- | --- | --- | --- |
| Intercept | -1.1 | 0.75 | 0.13 |
| Ln SARS-CoV-2 pMN IC50 | 0.16 | 0.06 | 0.0088 |
| Patient | 2.9 | 0.26 | <0.0001 |
| Ln NL63 pMN IC50 | 0.35 | 0.11 | 0.0018 |

When assessing the assumptions of the linear regression we found no trends in the residuals, no overly influential data points, and the qq-plot indicated that the assumption of normality was not violated. When assessing the assumption of proportional odds in our proportional odds logistic regression we used simple logistic regression and found indicators that the effect of predictor variables was not independent of COVID-19 severity. First the effect of the patient indicator was estimated with a great deal of uncertainty in the individual logistic regression models. However, this was expected due to perfect separation, i.e. all individuals with severity >4 were patients, and was not an issue in the full proportional odds model. The effect of SARS-CoV-2 pMN IC50 was estimated to be greater when distinguishing between severity < 4 and >= 4. This observation is most likely because severity 4 is the severity where patient status is least able to separate the groups.

Finally, and most importantly, the effect of NL63 neutralisation was slightly higher when distinguishing between severity 1 and higher. This suggests that the effect of NL63 neutralisation is greatest when distinguishing between asymptomatic and symptomatic cases. However, the estimated effect size was also positive for all other models. We reran our proportional odd logistic regression on the 67 samples from symptomatic cases and found that the effect of NL63 was only borderline significant (LR=3.5, df=1, p=0.06).

### Does SARS-CoV-2 exposure increase HCoV neutralisation?

To investigate if SARS-CoV-2 infection increases HCoV neutralisation we compared SARS-CoV-2 seronegative and SARS-CoV-2 seropositive sample neutralisation for NL63, HKU1, 229E, and for reference SARS-CoV-2 (figure 5). We found a small but significant 1.5-fold increase in geometric mean of NL63 neutralisation after accounting for the effects of sex and age (serostatus ß=0.43, SE=0.14, p=0.003; sex ß=-0.33, SE=0.14, p=0.018; age ß=0, SE=0.01, p=0.90). We found no difference in HCoV neutralisation between seropositive and seronegative samples for HKU1 (serostatus ß=-0.73, SE=0.50, p=0.143; sex ß=1.5, SE=0.49, p=0.002; age ß=0.03, SE=0.02, p=0.150) or 229E (serostatus ß=-0.01, SE=0.24, p=0.978; sex ß=0.39, SE=0.24, p=0.104; age ß=0.02, SE=0.01, p=0.021). As expected, seropositive samples showed a large increase in SARS-CoV-2 neutralisation (205-fold increase, p<0.001).

**Figure 5.**
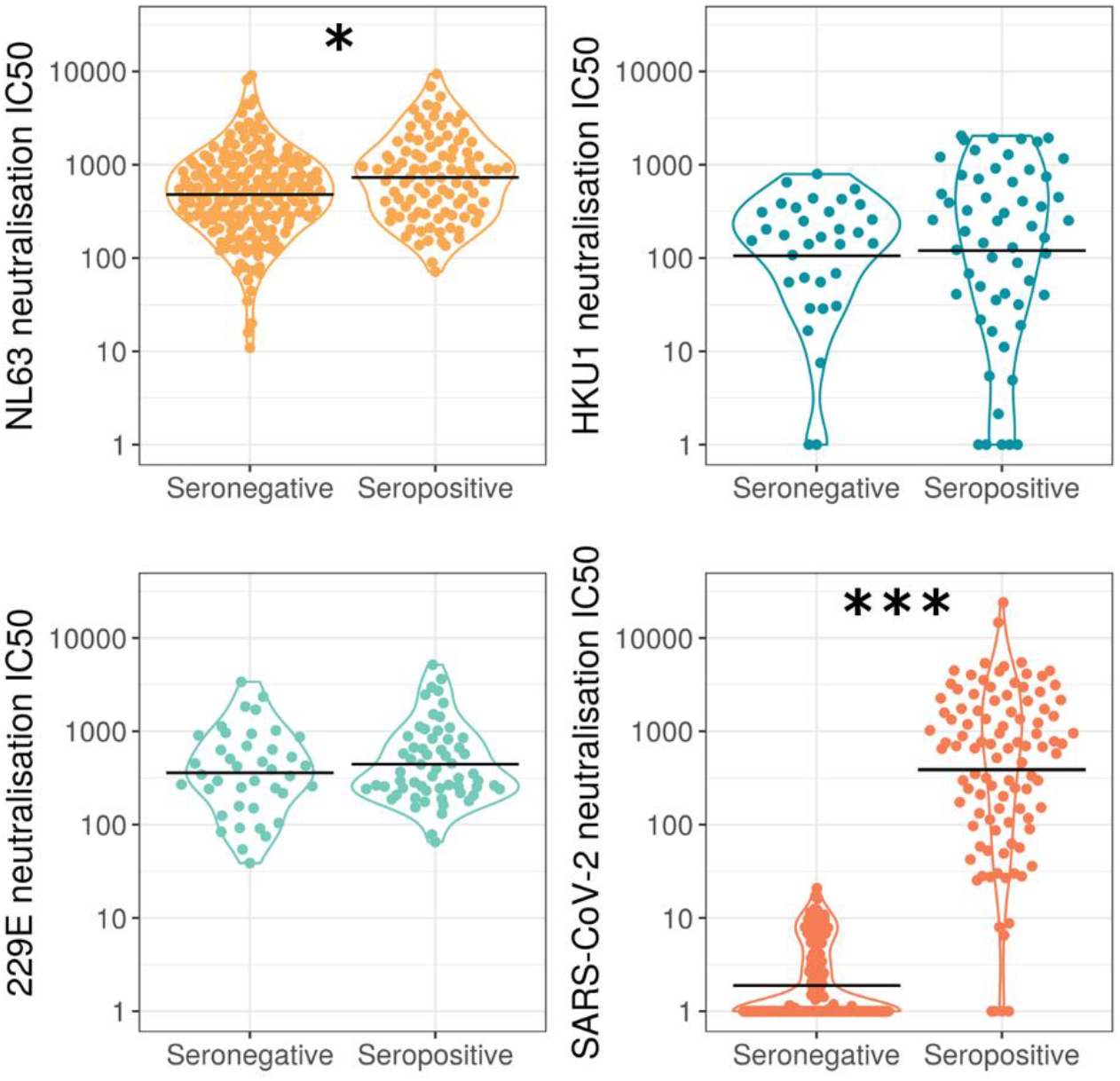
Comparing neutralisation of SARS-CoV-2 seropositive and seronegative samples revealed significant differences for SARS-CoV-2 (p=<0.001) and NL63 (p=0.018). However, we observed no significance for HKU1 (p=0.143) and 229E (p=0.978). Horizontal black lines indicate geometric means. Samples were grouped as seropositive or seronegative regardless of being from patients or HCWs.

We also found that SARS-CoV-2 vaccination significantly increased NL63 neutralisation (p=0.0001, figure 6). We used paired pre- and post-vaccination samples to identify a significant increase in NL63 neutralisation post-vaccination. The geometric mean of fold-increase in NL63 neutralisation, 2.2 was similar to the difference between seropositive and seronegative cases. Vaccination appears to cause a similar fold-increase in NL63 neutralisation to all samples.

**Figure 6.**
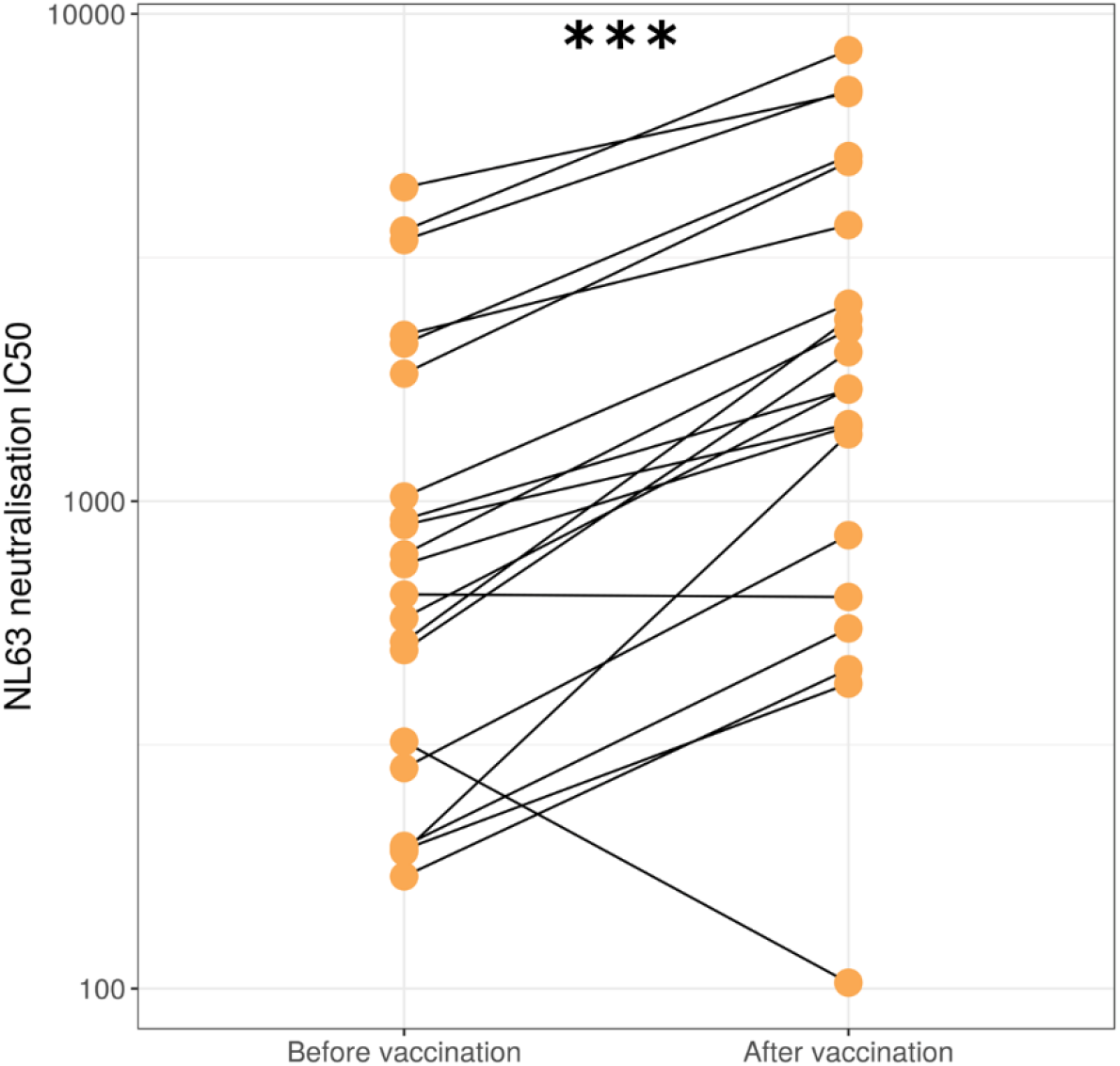
NL63 neutralisation in HCWs before and approximately one month after first dose of SARS-CoV-2 vaccination (p=<0.001).

### Correlations between neutralisation of different viruses and spike binding

We found no correlation between HCoV neutralisation and SARS-CoV-2 neutralisation in SARS-CoV-2 seropositive individuals (figure 7), suggesting that HCoV neutralising antibodies do not neutralise SARS-CoV-2. The Spearman’s rank correlation coefficient was non-significant for each HCoV tested, NL63 r=0.05, p=0.65, HKU1 r=0.1, p=0.48, 229E r=0.15, p=0.24. This is in keeping with our finding that HCoV neutralisation was not associated with lower COVID-19 severity.

**Figure 7.**
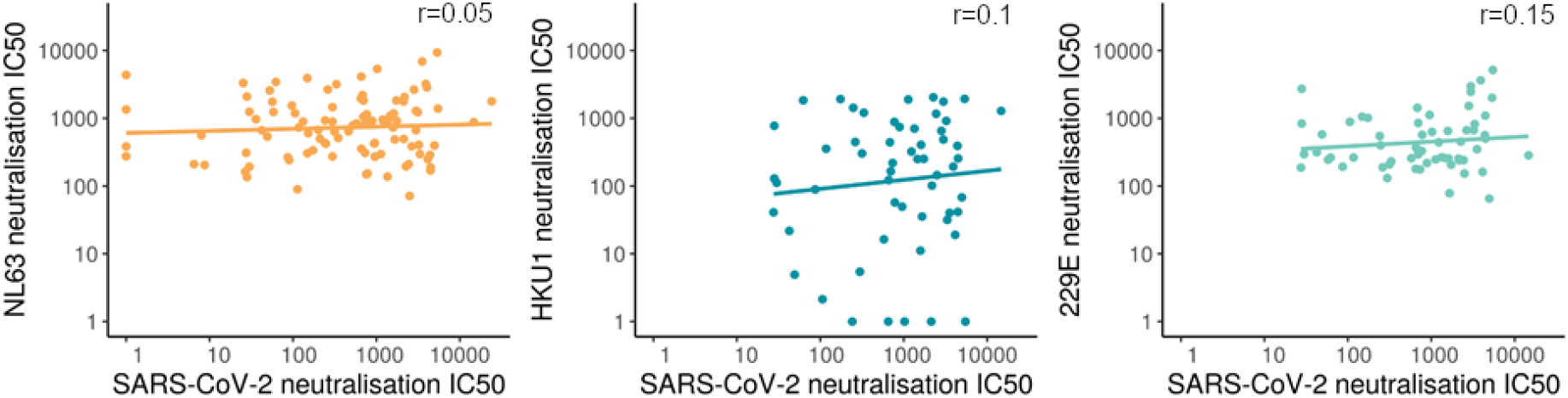
The relationship between SARS-CoV-2 neutralisation and HCoVs in SARS-CoV-2 seropositive samples with a linear line of best fit. Spearman’s rank test reveals no significant correlation for each HCoV tested.

Although SARS-CoV-2 spike binding was closely correlated with SARS-CoV-2 neutralisation, there was little correlation between spike binding and neutralisation for HCoVs. However, there was strong positive correlation between spike binding to the different HCoVs (figure 8).

**Figure 8.**
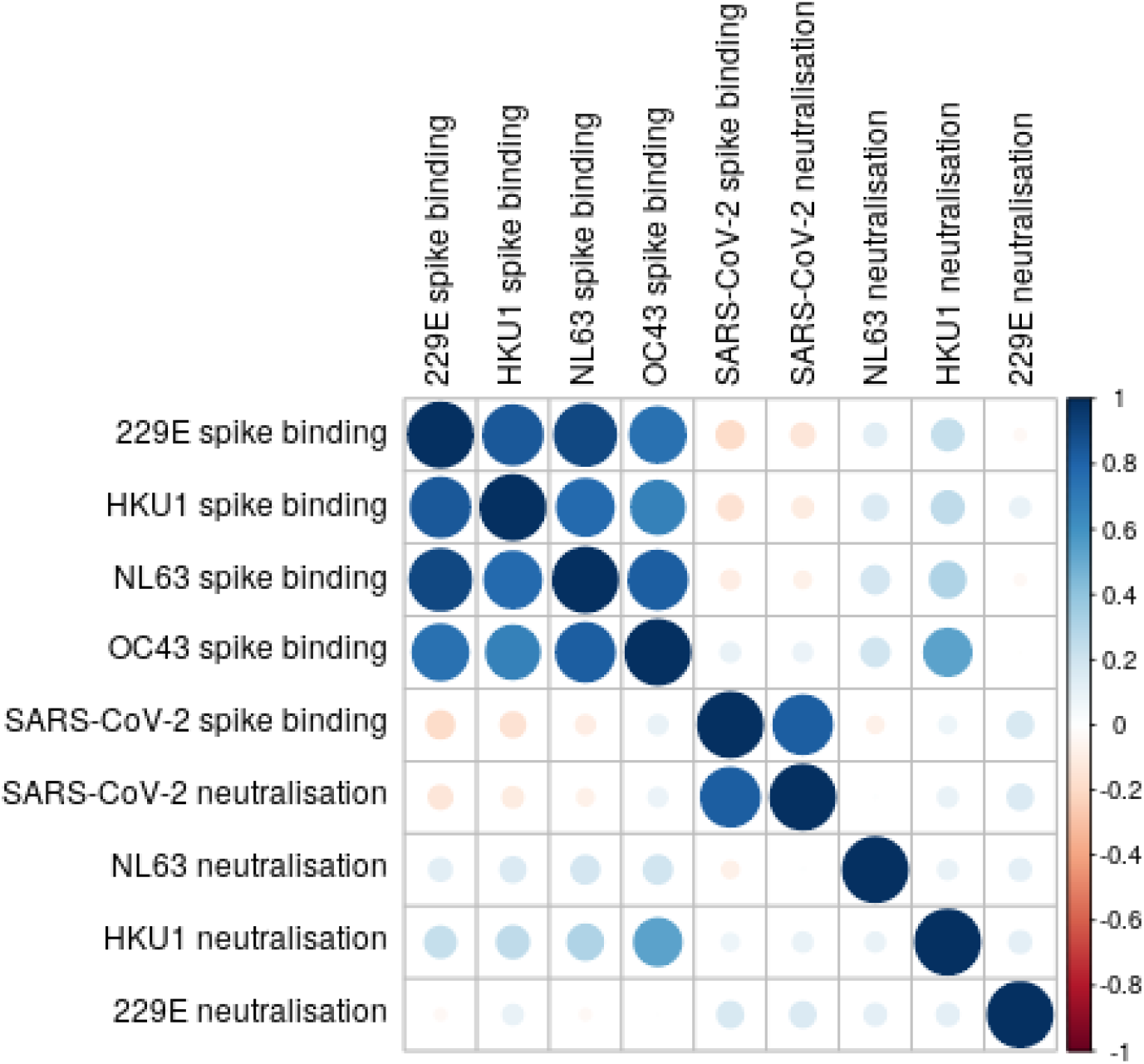
Correlation matrix of virus binding and neutralisation. Larger darker circles indicate stronger correlations, as measured by Spearman’s rank correlation coefficient. Blue circles indicate positive correlation and red circles indicate negative correlations

## Discussion

HCoVs cause frequent mild infections in humans with most people becoming infected during infancy (27), and reinfections remain a common occurrence after aproximately 12 months (28). Because of the frequency of HCoV infections much of the population will possess some immune response to one or more of the HCoVs. Therefore, it is important to identify any impact HCoV immune responses have on SARS-CoV-2, or the severity of COVID-19 disease it causes. We found that almost all samples tested had antibodies that neutralised NL63 and 229E, and more than three quarters of all samples had neutraling titres to HKU1. We found that HCoV neutralisation was not associated with protection from COVID-19. This result builds on previous work by Anderson et al. (2021) which showed that HCoV binding did not protect against COVID-19. Despite similar conclusions, we found that HCoV spike binding did not correlate well with HCoV neutralisation.

Given that NL63 was the HCoV most likely to influence COVID-19 severity because it also targets the ACE2 receptor for cell entry, we analysed a larger sample size for NL63 neutralisation. Interestingly, we found that NL63 neutralisation was positively correlated with COVID-19 severity after accounting for SARS-CoV-2 neutralisation using two separate statistical methods. This relationship is most clearly seen within HCW with mild disease, explaining why we did not find that patients had higher NL63 neutralisation. A previous report found that all samples with severe COVID-19 disease showed very low levels of NL63 neutralising antibodies (29). However, this contrasts with our findings which demonstrate a range of NL63 neutralisation titres in individuals with both high and low COVID-19 severity scores.

Here we present evidence that exposure to SARS-CoV-2 increases neutralisation of NL63 despite no cross neutralisation. We show that SARS-CoV-2 vaccination increases neutralisation of NL63 and that NL63 neutralisation was higher in SARS-CoV-2 seropositive samples. One limitation of this study is the lack of paired samples immediately before and after infection to measure the effect of SARS-CoV-2 infection on NL63 neutralisation directly. If moderate to severe COVID-19 disease causes an increase in NL63 neutralisation it would explain the observed relationship between COVID-19 severity and NL63 neutralisation. The increase in NL63 neutralisation after vaccination suggests the possibility that SARS-CoV-2 vaccination may also provide protection against the common cold caused by NL63.

Spike protein binding was highly correlated between the HCoVs, however; there was relatively little correlation between HCoVs and SARS-CoV-2. If the correlation between HCoV binding was driven by cross reactive antibodies we would also expect them to correlate with SARS-CoV-2 as it is more closely related to the betacoronavirus HCoVs than 229E and NL63 are. We interpret the lack of correlation in spike binding between SARS-CoV-2 as evidence that HCoV spike antibodies are likely not cross reactive but co-occurring, i.e. people who are exposed to one of the HCoVs are likely to be exposed to other HCoVs (28).

We found that SARS-CoV-2 neutralisation does not correlate with neutralisation of any HCoV we tested. At first this seems to contradict several studies reporting cross-reactive binding and neutralisation (7,8); however, these studies found only a very small proportion of people not exposed to SARS-CoV-2 displayed cross-reactive antibodies. Ng et al 2020 found that less than 1% of pre-pandemic samples showed SARS-CoV-2 RBD binding antibodies. This suggests that the majority of HCoV antibodies do not cross-react with SARS-CoV-2 and is in keeping with our results that HCoV neutralisation is not correlated with SARS-CoV-2 neutralisation, nor does it provide protection against COVID-19, which is consistent with a similar study (30). On the other hand, a study observed that a recent HCoV infection may provide some degree of protection (31). We do not have information on timing of HCoV infection so cannot test this relationship in this study.

One of the limitations of pseudotyped viruses is that they possess only the spike protein, therefore; the effects of other viral proteins remain in question. The nucleocapsid protein (N) shows highly conserved motifs in the N-terminus, observed across a wide range of the HCoVs and SARS-CoV-2 (32). Cross reactivity between SARS-CoV-1 N-antibodies and several animal coronaviruses were previously described, despite lack of cross reactive spike antibodies (33). Similarly, several reports have found cross-reactive antibodies between HCoVs and SARS-CoV-2 S, M and N proteins (7–9). Importantly, a report observed that N-antibodies of several viruses activate the TRIM21 pathway, which then drives cytotoxic T-cell activation (34). Given that SARS-CoV-2 reactive T-cells were detected in healthy donors (10,11,35) several studies have investigated T-cell activation between HCoVs and SARS-CoV-2, with differing reported outcomes. One group did not observe any N-specific T cells from a pool of healthy blood donors, despite finding S and M specific T cells (36). This contrasts with another study that identified CD4 and CD8 T cells that recognised multiple regions of the N protein (37), and another report that also detected an individual with T cells reactive to N (38). Further studies are required to ascertain the impact of the N-protein with regards to T cell activation.

## Data Availability

All data would be made available upon request

## Acknowledgements

This study was undertaken by the Humoral Immune Correlates to COVID-19 (HICC) consortium, funded by the UKRI and NIHR; grant number G107217 (COV0170 - HICC: Humoral Immune Correlates for COVID19). RW and SE received funding from the StMWK (ForCOVID, Bavaria, Germany). We thank the RPH Foundation Trust COVID-19 Research and Clinical teams for supporting recruitment to this study, HCWs and Outpatients who participated in this study.

